# Functional fine-mapping of the 9p21.3 cardiovascular disease locus implicates the lncRNA MIR31HG as an effector transcript in endothelial cells

**DOI:** 10.64898/2026.10.15.26365688

**Authors:** Artemy Zhigulev, Flore-Anne Poujade, Nikita Gryzunov, Andrey Buyan, Haonan Li, Otto Bergman, Minghui Liu, Sailendra Pradhananga, Anders Franco-Cereceda, Peter Swoboda, Ivan V. Kulakovskiy, Per Eriksson, Pelin Sahlén, Hanna M Björck

**Affiliations:** Cardiology Unit, Center for Molecular Medicine, Department of Medicine, Karolinska Institutet, Karolinska University Hospital, Stockholm, Sweden; Science for Life Laboratory, School of Engineering Sciences in Chemistry, Biotechnology and Health, Division of Gene Technology, KTH Royal Institute of Technology, Solna, Sweden; Institute of Protein Research, Russian Academy of Sciences, Pushchino, Russia; Life Improvement by Future Technologies (LIFT) Center, Moscow, Russia; Institute of Biomedical Problems, Russian Academy of Sciences, Moscow, Russia; Department of Medicine Huddinge (MedH), Biosciences and Nutrition Unit, Karolinska Institutet, Stockholm, Sweden; Department of Molecular Medicine and Surgery, Karolinska Institutet, Karolinska University Hospital, Stockholm, Sweden; Institute of Biochemistry and Genetics, Ufa Federal Research Centre of the Russian Academy of Sciences, Ufa, Russia; Vavilov Institute of General Genetics, Russian Academy of Sciences, Moscow, Russia

**Keywords:** Cardiovascular disease, Coronary artery disease, 9p21.3, lncRNA, MIR31HG, HiCap, chromatin interactions, enhancer-promoter regulation, endothelial cells

## Abstract

**BACKGROUND:** The chromosome 9p21.3 locus is among the strongest genetic risk loci for cardiovascular disease (CVD), yet the regulatory mechanisms underlying this association remain poorly understood. Previous vascular disease studies focused predominantly on the role of this locus in smooth muscle cell (SMC) dysfunction, whereas its potential role in endothelial cells (ECs) remains less well characterized. Here, we sought to identify candidate regulatory variants at 9p21.3, their distal target genes, and effector transcripts that contribute to CVD-related phenotypes in ECs.

**METHODS:** Promoter capture Hi-C was used to map chromatin interactions at 9p21.3 in primary human ascending aortic SMCs (n=6), supplementing the previously obtained ECs (n=16) dataset. To support the regulatory relevance of these interactions, expression quantitative trait locus analysis was performed in 136 vascular biopsies. Variant effects were prioritized *in silico* using AlphaGenome. The regulatory activity of the prioritized region was validated using CRISPR activation.

**RESULTS:** Chromatin interaction mapping identified a complex regulatory architecture at the 9p21.3 locus involving 19 putative regulatory elements and 15 genes across ECs and SMCs. *MIR31HG* showed the most recurrent distal promoter connectivity with the 9p21.3 region. Consistent with the chromatin interaction map, expression of *MIR31HG* transcripts was associated with genetic variation at the 9p21.3 locus. In ECs, the lncRNA MIR31HG, but not miR-31, was responsive to hemodynamic flow. Depletion of *MIR31HG* in ECs was associated with endothelial activation. Epigenetic tracks and deep learning predictions highlighted an active enhancer element at 9p21.3, which was subsequently validated by CRISPR activation and increased *MIR31HG* expression, confirming its capacity to regulate *MIR31HG* over a large genomic distance of approximately 538 kb.

**CONCLUSIONS:** Altogether, these data implicate lncRNA MIR31HG as a likely effector of the 9p21.3 CVD locus, linking the cis-regulatory element and heritable genetic variation in the respective region to endothelial activation through long-range enhancer regulation.

**Clinical Perspective:** **What Is New?**

- Promoter capture Hi-C in primary human vascular cells identified recurrent long-range interactions between the 9p21.3 cardiovascular disease locus and the *MIR31HG* promoter, extending the regulatory landscape beyond the nearest genes.
- Expression of *MIR31HG* transcripts was associated with 9p21.3 genetic variation in vascular tissue; the lncRNA MIR31HG was responsive to hemodynamic flow in endothelial cells and influenced the endothelial activation transcriptional programs upon depletion.
- Integrated sequence-based, epigenomic, genetic, and CRISPR activation analyses nominated the 9p21.3 enhancer as a regulator of *MIR31HG* in endothelial cells.

**What Are the Clinical Implications?**

- The findings suggest that the cardiovascular disease associations of 9p21.3 may involve stronger than previously anticipated endothelial regulatory mechanisms in addition to characterized effects in other vascular cell types.
- lncRNA MIR31HG provides a molecular link between noncoding genetic variation at 9p21.3 and endothelial cell-state regulation relevant to vascular disease.

## Introduction

Cardiovascular disease (CVD) remains a leading cause of morbidity and mortality worldwide.^1^ Genome-wide association studies (GWAS) have identified numerous loci associated with CVD risk, but translating these genetic associations into molecular mechanisms remains a major challenge.^2^ Among the loci identified to date, chromosome 9p21 stands out as one of the strongest and most consistently replicated genetic risk loci for CVD, and fine-mapping of the region identified the 9p21.3 locus, a 58-kb linkage disequilibrium block overlapping the long noncoding RNA (lncRNA) gene *ANRIL*.^3–8^ Consequently, considerable efforts have focused on determining whether *ANRIL* and its nearest genes mediate the CVD risk associated with this locus.^9–11^ However, despite more than a decade of investigation, the mechanism through which genetic variation at 9p21.3 contributes to CVD remains incompletely understood.

Disease-associated noncoding variants may act through enhancer-promoter interactions; however, they target the nearest gene only in 30% of cases.^12,13^ In fact, increasing evidence indicates that they can regulate genes over large genomic distances, raising the possibility that the critical effectors of 9p21.3 extend beyond genes located immediately adjacent to the risk interval.^14^ Genetic variation within enhancers can alter transcription factor (TF) binding, thereby affecting chromatin accessibility, enhancer activity, and finally reshaping transcriptional programs and contributing to disease susceptibility.^15^ Advances in chromatin conformation mapping have therefore provided new opportunities to connect disease-associated noncoding variants with their distal target genes and to identify the regulatory pathways through which genetic risk is mediated.^16–19^

Most functional studies of 9p21.3 in vascular disease have focused on vascular smooth muscle cells (SMCs), while the contribution of endothelial cells (ECs) is less well characterized. Here, we analyzed promoter-centered chromatin interactions at 9p21.3 in primary human ascending aortic ECs and SMCs to establish previously unrecognized EC effectors. We identified recurrent long-range interactions with the *MIR31HG* promoter and examined the relationship between 9p21.3 genotype and the expression of *MIR31HG* transcripts in human vascular tissues. We further characterized transcript responses to hemodynamic flow and determined the transcriptional consequences of *MIR31HG* depletion in ECs. Finally, we integrated endothelial chromatin features and sequence-based variant prioritization, including TF motif analysis, to identify the candidate regulatory region and tested its ability to activate *MIR31HG* using CRISPR activation (CRISPRa).

Together, our study identifies an endothelial regulatory axis involving lncRNA MIR31HG and provides functional support for long-range regulation of this transcript by an enhancer within the 9p21.3 cardiovascular disease locus.

## Methods

### Ethics

The study was approved by the Human Research Ethics Committee at Karolinska Institutet (application numbers 2006/784−31/1 and 2012/1633−31/4), Stockholm, Sweden. Written informed consent was obtained from all individuals in accordance with the Declaration of Helsinki. All methods were carried out following relevant guidelines.

### Patient cohorts

In this study, we included patients (n=22) undergoing elective open-heart surgery for ascending aortic dilatation (diameter > 40 mm), with or without concomitant aortic valve replacement, at the Cardiothoracic Surgery Unit, Karolinska University Hospital, Stockholm, Sweden. None of the individuals had significant coronary artery disease, based on coronary angiography.^20^ Participation in this project is entirely voluntary and does not provide financial compensation. Patients used to obtain ECs (n=16) were described in our previous study.^17^ Briefly, the gender distribution was 7 males / 9 females; ages varied from 47 to 80 years, with an average of 66 years, and all patients were Caucasian except one. Patients used to obtain SMCs (n=6) had a gender distribution of 5 males / 1 female; ages varied from 45 to 78 years, with an average of 61 years, and all patients were Caucasian except one.

### Cell cultures

Primary aortic smooth muscle cells were enzymatically isolated from the ascending aorta of patients. They were grown in SmBM basal medium supplemented with the SMGM-2 Bullet Kit (CC-3182, Lonza) in T75 flasks and passaged when they reached 80% confluency. Cells were used no later than P6. Constant environmental conditions of 37 °C with 5% CO_2_ were maintained throughout the growth of all cells.

EA.hy926 cells were purchased from ATCC (CRL-2922) and grown in DMEM high glucose (32430027, Gibco) supplemented with 10% fetal bovine serum and 1% penicillin-streptomycin. Cells were passaged when reaching 80% confluency and used up to P10.

### Promoter capture Hi-C

High-throughput chromosome conformation capture followed by targeted sequence capture (HiCap) was performed as previously described.^21^ Briefly, the method starts with cross-linking the chromatin with 1% formaldehyde, followed by digesting DNA with the 4-cutter FastDigest MboI (↓GATC; ThermoFisher Scientific) into ∼700 bp fragments. Spatially close fragments are then ligated with T4 DNA ligase (New England Biolabs) for 4.5 h at 16 °C. Finally, promoter-enhancer interactions are captured using a custom probe set SureSelect XT Target Enrichment System (Agilent). It included 49,927 probes targeting 23,111 promoters (of 19,469 genes), 1123 GWAS variants associated with cardiovascular disease and lipid traits, and 2408 negative control regions with no known regulatory activity.

The resulting enriched DNA libraries were sequenced in-house via Illumina single-index paired-end sequencing on the NextSeq 500 platform (Illumina Inc.). HiCap libraries were mapped to hg38 using bwa-mem2 (version 2.2.1-20211213-edc703f) and PairTools (version 1.0.2).^22,23^ HiCapTools v1.3.2 was used to call statistically significant interactions in all samples (five or more read pairs supporting each interaction and a Bonferroni-adjusted *P* < 0.05).^24^ We further filtered out trans interactions, long-range interactions with an interaction distance higher than or equal to 5 Mb, and DpnII fragments with lengths higher than or equal to 7500 bp.

### Expression quantitative trait locus analysis

Expression quantitative trait locus (eQTL) analysis was based on the previously published dataset.^25^ It was performed to assess associations between the expression of 15 candidate genes and genotype dosage (0, 1, or 2 alternate alleles) within the predefined genomic region. The genomic region was defined as the 58 kb LD block identified from the literature (chr9:22062302:22120390), flanked by 7500 bp windows. For each SNP–gene pair, linear regression was used to estimate the allelic effect, and the multiple testing correction was performed using the Benjamini–Hochberg false discovery rate (FDR) procedure.

### *In vitro* flow exposure and gene expression measurements

*In vitro* flow exposure was performed as previously described.^26^ Briefly, EA.hy926 cells were plated on gelatine-coated Teflon-bordered cell culture slides (75×25×10mm, Flexcell International Corp.) and cultured for 40 h with 5% CO_2_ at 37 °C. Culture slides were then inserted into a parallel-plate Streamer device (Flexcell International Corp., Hillsborough, NC, US) and exposed to either bidirectional oscillatory shear stress of ±12 dynes/cm^2^ or unidirectional laminar flow of 12 dynes/cm^2^ for 48 h at 37 °C in 5% CO_2_. A Masterflex L/S peristaltic pump generated flow, and oscillation frequency was controlled by the Osci-Flow flow controller (Flexcell International Corp.). Matching EA.hy926 cells cultured under static conditions for the same period were used as controls. Total RNA was extracted using the miRNeasy Mini kit (Qiagen). RNA samples were hybridized and further processed at the core facility for Bioinformatics and Expression Analysis at the Department of Biosciences and Nutrition, NEO, Karolinska Institute, Huddinge, using the Human Transcriptome Array 2.0 (Affymetrix) and according to the manufacturer’s instructions.

### siRNA knockdown and RNA-seq analysis

Knockdown (KD) was performed on EA.hy926 cells using either siRNA against *MIR31HG* transcripts or control siRNA. The KD efficiency was validated using qPCR. Following KD, total RNA libraries were sequenced on NovaSeq6000. EdgeR was used for downstream analysis.^27^ The details are provided in Supplementary Methods.

### CRISPR activation

CRISPRa was performed using the dCas9-VP192 transcriptional activator together with guide RNAs (gRNAs) targeting the *MIR31HG* promoter and a distal candidate regulatory region. gRNAs were designed using the Benchling CRISPR design tool and selected based on their genomic positions, predicted specificity, and predicted activity. Three gRNAs were selected for each target region and pooled for CRISPRa experiments.

gRNA transcriptional units were generated by PCR assembly as previously described.^28^ Briefly, individual gRNA expression cassettes were assembled from a U6 promoter fragment, a guide-specific oligonucleotide containing the target sequence, and a common scaffold/terminator fragment using high-fidelity PCR. The resulting gRNA transcriptional units were purified and pooled before transfection.

Pooled gRNA transcriptional units were co-transfected with the dCas9-VP192 expression construct into hTERT-RPE1 cells. The *MIR31HG* promoter and distal regulatory region were targeted individually or in combination. Following CRISPRa, total RNA was isolated and the expression of *MIR31HG* transcripts was quantified by RT-qPCR.

### Motif activity response analysis

Gene-level log_2_CPM expression estimates derived from the *MIR31HG* KD RNA-seq data were supplied to the motif activity response analysis, MARA, with MARAdoner to identify key TFs with altered activity in knockdown cells.^29,30^ To locate the heart-specific transcription start sites, we used the CAGE-Seq heart atlas.^31^ The details are provided in Supplementary Methods.

### Variant effects annotation with AlphaGenome

To predict cell type-specific variant effects on enhancer activity, we employed AlphaGenome, a multimodal sequence-to-function model for predicting regulatory activity and estimating the effects of genetic variants.^32^ Briefly, first, we selected a subset of AlphaGenome tracks representing active chromatin (CAGE signal, DNase accessibility, and key histone modifications) in relevant cell types. Second, to recalibrate the predicted scores using an empirical background distribution, we assembled the control set of 10,000 genomic windows anchored at common SNPs selected within ENCODE peaks matching the corresponding AlphaGenome tracks. Finally, for each assay and cell type, we summarized the predictions as the mean predicted activity and the mean variant effect estimates for each group of tracks (e.g., H3K27ac in epithelial cells) and rescaled the scores against the background distribution, i.e., replaced each value by the respective background quantile, making the scores comparable across different tracks and cell types. In this analysis, we used the GRCh38 human genome assembly. Technical details are provided in Supplementary Methods.

## Results

### Chromatin interaction mapping identifies *MIR31HG* as a target gene of the 9p21.3 locus

To characterize the regulatory landscape of the 9p21.3 CVD susceptibility locus, we integrated newly obtained promoter-centric chromatin interactome data of primary ascending aortic SMCs (n=6) with our previously published data of primary ascending aortic ECs (n=16). This way, we revealed an extensive interaction network comprising 19 putative regulatory elements and 15 genes across both cell types, with an average of 4.8 ± 2.6 promoter interactions per element (Figure 1A-F).

**Figure 1.**
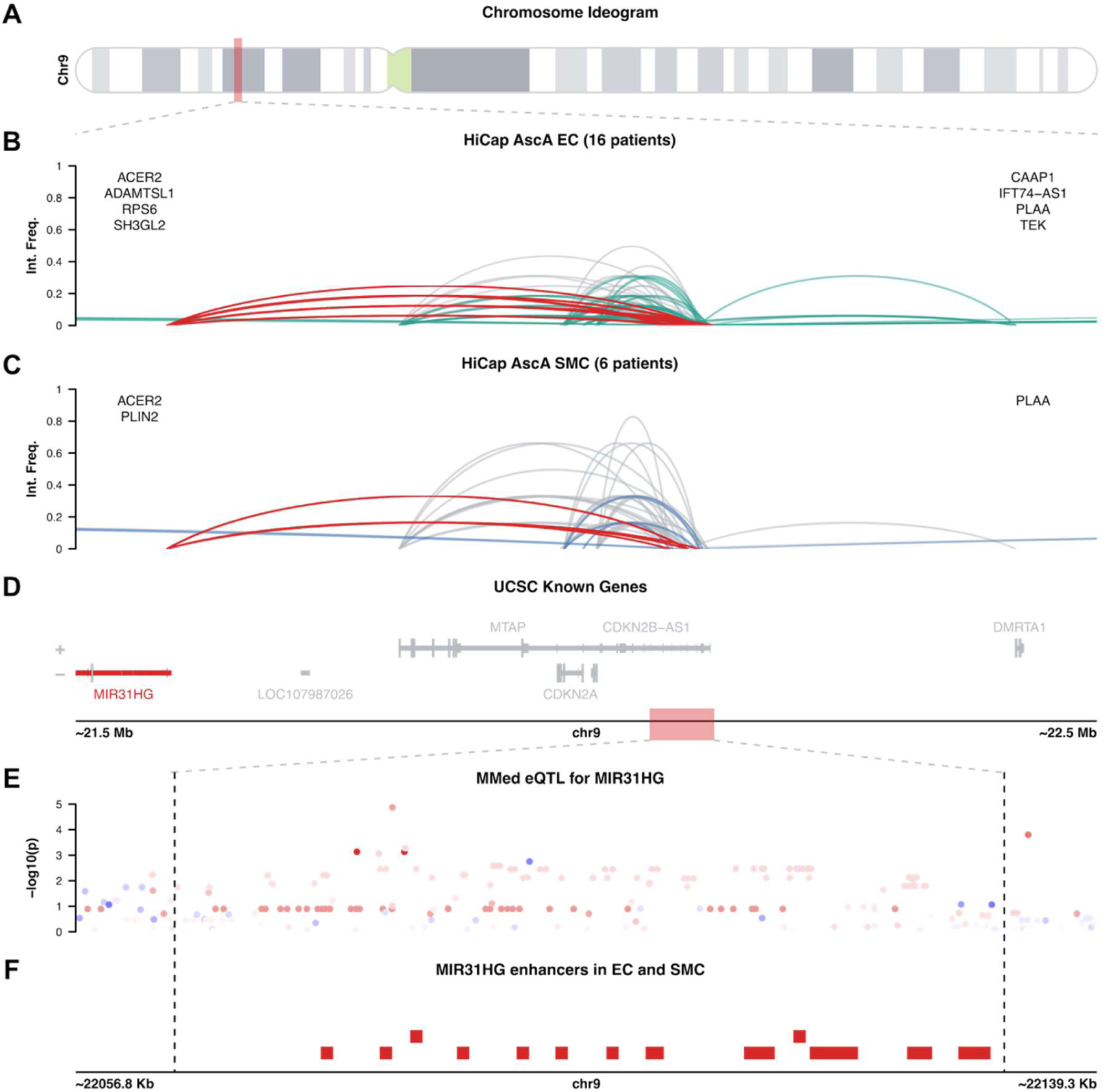
Chromatin interaction landscape of the 9p21.3 cardiovascular disease risk locus. **A,** Chromosome 9 Ideogram. Enhancer-Promoter interaction maps in **(B)** ascending aorta ECs and **(C)** SMCs (read count > 4, Bonferroni-adjusted *P* < 0.05). Grey lines indicate interactions shared between cell types, whereas colored lines indicate cell-type-specific interactions. Genes flanking the interaction maps indicate additional long-range targets of the 9p21.3 locus. **D,** UCSC Known Genes map. **E,** eQTL associations between the *MIR31HG* promoter and genetic variation at9p21.3 in internal thoracic artery intima-media samples (assessed by linear regression using allelic dosage). Color indicates the direction and magnitude of the effect size. **F,** HiCap-derived enhancers interacting with the *MIR31HG* promoter. AscA, ascending aorta; ECs, endothelial cells; eQTL, expression quantitative trait locus; HiCap, promoter capture Hi-C; MMed, internal thoracic (mammary) artery; SMCs, smooth muscle cells.

The overall interaction landscape was substantially shared between ECs and SMCs (Figure 1B and 1C). Consistent with previous studies, the strongest and most recurrent interactions involved the neighboring genes *CDKN2A*, *CDKN2B*, and *MTAP* (Table 1). However, *CDKN2B-AS1* (also known as *ANRIL*) showed limited connectivity with the locus.

**Table 1.** Genes interacting with 9p21.3 based on HiCap.

| Genes | HPO, scRNA-seq, CPM |  | Interactions with the GWAS SNPs in the 9p21.3 locus, % |  | Distance, Kb | Groups |
| --- | --- | --- | --- | --- | --- | --- |
|  | EC | SMC | EC | SMC |  |  |
| CDKN2B | 3.6 | 5.6 | 15.3 | 24.3 | 89 | Classical genes |
| CDKN2B-AS1 | NA | NA | 6.9 | 2.7 | 103 |  |
| CDKN2A | 1.6 | 1.7 | 19.4 | 27 | 123 |  |
| MTAP | 2.1 | 3.5 | 16.7 | 21.6 | 295 |  |
| DMRTA1 | 0.5 | 0.4 | 5.6 | 2.7 | 349 | Novel closest genes |
| **MIR31HG** | NA | NA | 13.9 | 10.8 | 538 |  |
| ACER2 | 62.7 | 20.6 | 1.4 | 2.7 | 2689 | Other |
| RPS6 | 2823 | 2238.9 | 1.4 | 0 | 2718 |  |
| PLIN2 | 83.9 | 28.2 | 0 | 2.7 | 2971 |  |
| ADAMTSL1 | 109.9 | 49.1 | 1.4 | 0 | 3624 |  |
| SH3GL2 | 1.9 | 2.2 | 1.4 | 0 | 4519 |  |
| CAAP1 | 32.8 | 34.6 | 2.8 | 0 | 4795 |  |
| PLAA | 36.8 | 40.9 | 6.9 | 5.4 | 4849 |  |
| IFT74-AS1 | NA | NA | 5.6 | 0 | 4858 |  |
| TEK | 321.2 | 3.9 | 1.4 | 0 | 5011 |  |

Importantly, the interaction landscape extended beyond genes immediately adjacent to 9p21.3. Significant long-range interactions were detected with genes located approximately 0.35 −5 Mb from the risk locus, including *MIR31HG*, *DMRTA1*, *ACER2*, *RPS6*, *PLIN2*, *ADAMTSL1*, *SH3GL2*, *CAAP1*, *PLAA*, *IFT74-AS1*, and *TEK*. Among these genes, *MIR31HG* showed the highest frequency of interactions across both ECs and SMCs, emerging as the most prominent distal target.

We next examined whether the evidence of chromatin interaction was supported by genetic regulation of candidate target genes. Expression quantitative trait locus (eQTL) analysis was performed using expression measurements in 49 aortic intima-media samples (AMed), 43 aortic adventitia samples (AAdv), and 44 internal thoracic artery intima-media (mammary) samples (MMed) (Table S1). The strongest association was observed between the expression of *MIR31HG* transcripts and genetic variation at 9p21.3 in MMed (beta = 0.76, FDR < 0.05; Figure 1E). Although after the multiple testing correction, there was insufficient statistical power (FDR > 0.05, *P* < 0.05) to support associations of the locus with *ADAMTSL1* in MMed (beta = −0.78), *ACER2* in AMed (beta = −0.43) and *TEK* in AAdv (beta = −0.46), these genes may still represent interesting cases for further investigation (Figure S1). No significant associations were detected for *CDKN2A*, *CDKN2B*, or *MTAP* in these datasets. These findings provide complementary chromatin and genetic evidence that *MIR31HG* is a candidate distal target of the 9p21.3 locus in vascular tissue.

### lncRNA MIR31HG, but not miR-31, is flow-responsive in endothelial cells

*MIR31HG* gives rise to both the lncRNA MIR31HG and microRNA (miRNA) miR-31 from a shared promoter. Although miR-31 has previously been implicated in vascular biology, its reported functions have largely focused on SMCs, whereas the role of lncRNA MIR31HG in vascular cells remains poorly characterized. Given the recurrent interaction profile between 9p21.3 regulatory elements and the *MIR31HG* promoter in ECs, we sought to determine whether lncRNA MIR31HG represents a previously unrecognized effector of the 9p21.3 locus in ECs. Because atheroprone regions are exposed to distinct hemodynamic forces, we hypothesized that a disease-relevant effector transcript may exhibit flow-responsive regulation.

We exposed ECs to laminar or oscillatory flow and measured the expression of lncRNA MIR31HG and miR-31 (Table S2). lncRNA MIR31HG expression was significantly altered under both laminar (effect size = 0.76, FDR=0.005, two-sided Wilcoxon rank-sum test) and oscillatory (effect size = 0.64, FDR=0.024, two-sided Wilcoxon rank-sum test) flow compared with static conditions (Figure 2A, left). MiR-31 expression remained unchanged across flow conditions (Figure 2A, right). This finding indicates that the functions of lncRNA MIR31HG extend beyond hosting miR-31, making it a more likely effector in ECs.

**Figure 2.**
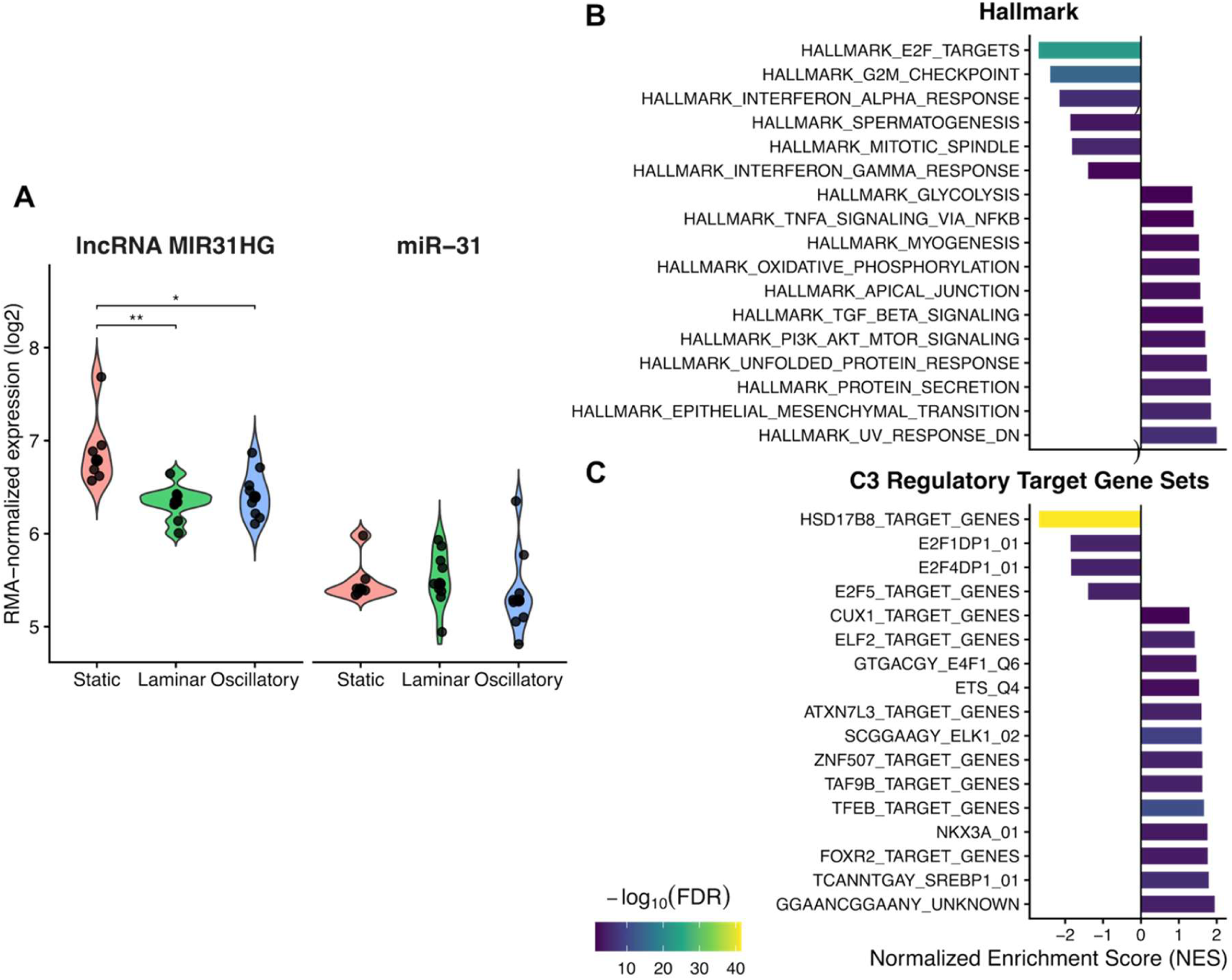
The role of *MIR31HG* in endothelial cells. **A,** Flow effects of the *MIR31HG* lncRNA, but not miR-31. Static conditions, n=6; laminar and oscillatory flow conditions, n=9. Two-sided Wilcoxon rank-sum tests were performed. * indicates FDR < 0.05; ** indicates FDR < 0.005. Gene set enrichment analysis (GSEA) of the transcriptomic profile following *MIR31HG* KD, showing enrichment results for **(B)** Hallmark gene sets and **(C)** Regulatory Target Gene Sets. FDR, false discovery rate; GSEA, gene set enrichment analysis; KD, knockdown; lncRNA, long non-coding RNA.

### Depletion of *MIR31HG* promotes endothelial activation transcriptional program

To investigate the functional consequences of *MIR31HG* loss, we performed siRNA-mediated knockdown (KD) in ECs using two independent oligonucleotides across five biological replicates (Figure S2A-C, Table S3). RNA-seq profiling detected broad transcriptomic changes with 578 differentially expressed genes (DEGs; abs(logFC) > 0.58, FDR < 0.05).

Gene set enrichment analysis (GSEA) revealed a transcriptional program consistent with endothelial activation (Figure 2B, Table S4). Genes associated with proliferative activity were downregulated, including E2F targets, G2/M checkpoint, and mitotic spindle genes, whereas genes involved in cellular transformation and remodeling programs, including epithelial-to-mesenchymal transition (EMT), TGF-β signaling, and apical junctional complex pathways, were upregulated. *MIR31HG* KD was also associated with the bidirectional changes of inflammatory signaling and upregulation of metabolic pathways.

To identify key TFs driving the transcriptomic changes, including potential E2F involvement, we performed motif activity response analysis (MARA; Table S5). This analysis predicted several TF families with differential activity upon *MIR31HG* KD (FDR < 0.1): reduced E2F (z-score=−5.4) and CUX/ONECUT (z-score=−4.0) activities and increased ETS activity (z-score=4.7) in agreement with GSEA performed against MSigDB TF targets set (FDR < 0.05; Figure 2C, Table S6). We then used RNA-seq expression patterns to prioritize members of these TF families relevant in the EC context (Figure S3). Among the DEGs whose expression was most strongly supported by the differential activity of these TF families were *CALU*, *SIK3*, *TRMT112*, and *CFP* (MARADONER Bayesian belief > 0.70, motif *P* < 0.005), which were also shown to play a role in CVDs.^33–35^

Together, these findings show that *MIR31HG* coordinates vascular endothelial changes through a specific set of TF families, consistent with the endothelial activation phenotype observed during cardiovascular disease.

### 9p21.3 locus harbors a distal enhancer and putative regulatory SNPs of *MIR31HG*

We next sought to identify candidate regulatory regions and respective sequence variants that likely contribute to the observed association between 9p21.3 and *MIR31HG*. We integrated experimentally measured endothelial H3K27ac chromatin data with enhancer predictions from the ENCODE-rE2G framework and 110 variants from the eQTL dataset, identifying a high-confidence putative enhancer region.^17,36^ To test whether this region could indeed affect *MIR31HG* transcription, we performed CRISPRa of the candidate enhancer (Figure 3A). Indeed, activation of the region located approximately 538 kb from the *MIR31HG* promoter significantly increased *MIR31HG* expression. Co-activation of the candidate enhancer and the *MIR31HG* promoter further increased its expression. These data confirm that the prioritized region acts as a long-range enhancer sufficient to upregulate *MIR31HG*.

**Figure 3.**
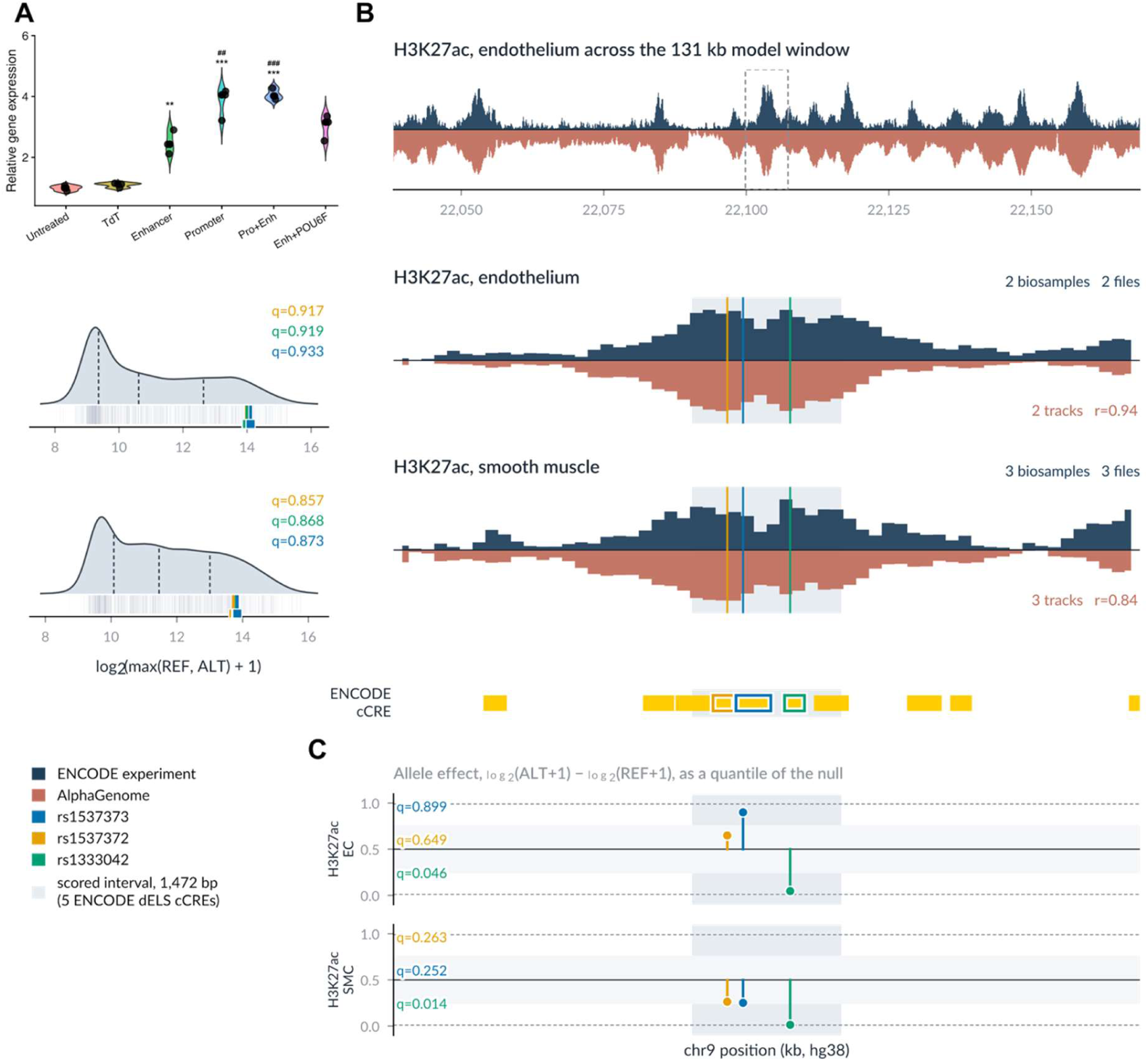
Gene regulation and genetic dissection of the *MIR31HG* enhancer. **A,** *MIR31HG* transcript levels in ECs measured by qRT-PCR 24 h after CRISPRa. Data represent *n*=3 biological replicates, with *n*=2 technical replicates per biological replicate. Statistical significance was assessed using one-way ANOVA followed by Tukey’s multiple-comparisons test. Asterisks indicate significant differences between the untreated condition and the other conditions (**FDR < 0.005; ***FDR < 0.0001). Hash symbols indicate significant differences between the enhancer-activated condition and the other conditions (##FDR < 0.005; ###FDR < 0.0001). **B,** H3K27ac profiles of the *MIR31HG* enhancer in ECs and SMCs. Blue profiles represent experimental ChIP-seq data, whereas brown profiles represent AlphaGenome predictions. ENCODE cCREs indicate candidate enhancers. **C,** AlphaGenome-predicted effect sizes differences between individual alleles. cCRE, candidate cis-regulatory element; CRISPRa, CRISPR activation; ECs, endothelial cells; FDR, false discovery rate; SMCs, smooth muscle cells.

Considering the genetic variation, across the 110 eQTL variants located within the 9p21.3 risk interval, only 45 overlapped putative regulatory elements interacting with the *MIR31HG* promoter (39 SNPs and 6 indels), and only three SNPs (rs1537372, rs1537373, and rs1333042) were located directly in the validated enhancer. To assess the regulatory impact of the SNPs, we employed AlphaGenome and predicted CAGE-seq, H3K27ac, H3K27me3, and H3K4me1 profiles, as well as variant effects for ECs and SMCs. Raw AlphaGenome scores for the enhancer regions were in good agreement with the experimental tracks (Figure 3B, Table S7). Moreover, loci harboring SNPs within the validated region were predicted to be more active in ECs than in SMCs.

Overall, we identified 20 SNPs within *MIR31HG*-interacting regions with predicted high allelic differences between the reference and alternative alleles (q<0.1 or q>0.9; Figure 3C, Table S8). While rs1333042 showed differential activity towards the reference allele in SMCs, its directionality did not agree with the eQTL results. In contrast, the allelic effect of rs1537373 in ECs was concordant with the eQTL: the risk allele G was associated with higher *MIR31HG* expression in the eQTL analysis, consistent with the AlphaGenome-predicted higher regulatory activity of the risk allele, although the predicted effect was of limited magnitude. Focused sequence motif analysis with PERFECTOS-APE suggests SNP-induced alteration of overlapping putative binding sites of the thyroid hormone receptor-related motifs, including RARA (fold change = 8.75) and NR1D1 (fold change = 1.32).^37^ Interestingly, the predicted binding affinity of NR1D1 is decreased on the alternative allele, aligning well with the allelic effect and the typical role of NR1D1 as a transcriptional repressor.^38^ Based on the Human Protein Atlas scRNA-seq dataset, both *NR1D1* (16 CPM in ECs vs. 9 CPM in SMCs) and *RARA* (40.5 CPM in ECs vs. 31.1 CPM in SMCs) are more highly expressed in vascular ECs compared to SMCs.^39^ However, the lack of supporting ChIP-Seq data does not allow for solidifying this conclusion. Altogether, these observations nominate rs1537373 as a candidate causal variant associated with *MIR31HG* expression in ECs.

## Discussion

The chromosome 9p21.3 locus remains one of the most extensively replicated genetic risk loci for CVD, yet the molecular mechanisms linking the genetic variation within this region to vascular disease remain incompletely resolved. As the CVD-associated variants often map to a noncoding genomic region, dysregulated gene expression rather than alterations in protein sequence is likely the driver of disease risk. In the present study, we mapped chromatin interactions in endothelial and smooth muscle cells isolated from ascending aortic biopsies of patients undergoing open-heart surgery, revealing a dense regulatory network of 15 genes and 19 putative regulatory elements.

Most previous work interrogating the cardiovascular risk conferred by the 9p21.3 locus has focused on *ANRIL* and neighboring genes (*CDKN2A*, *CDKN2B*, and *MTAP*). Indeed, we could show that the most recurrent interactions involved *CDKN2A*, *CDKN2B*, and *MTAP*, but *ANRIL* itself showed limited connectivity with risk-associated regulatory elements. Interestingly, our data also showed that the regulatory influence of 9p21.3 extended well beyond its immediate genomic neighborhood, consistent with growing evidence showing that disease-associated noncoding variants frequently exert their effects through long-range chromatin interactions rather than through the nearest gene. This underscores the importance of three-dimensional genome architecture in the interpretation of GWAS signals.

Notably, enhancer-gene predictions based on the ENCODE-rE2G model identified only three putative targets of the 9p21.3 locus: the two nearest proximal genes *CDKN2A* and *CDKN2B*, and the nearest distal gene *DMRTA1*, located approximately 350 kb from the locus, which is not expressed in vascular cells. In line with the previous evidence of the high inter-individual variation in TF binding and enhancer-promoter communication patterns among humans, these results further highlight the importance of individual-level analysis of the three-dimensional genome architecture.^17,40,41^

Among the 15 identified genes, *MIR31HG* emerged as the most prominent target gene, displaying recurrent interactions across both ECs and SMCs. To further support the 9p21.3 locus association with *MIR31HG*, we performed eQTL analysis in different types of intima-media (ECs+SMCs) and adventitia (fibrotic) tissues. Even with the limited sample size, we demonstrated that *MIR31HG* showed a stable signal in the intima-medial layer of the internal thoracic artery.

*MIR31HG* is particularly interesting as its promoter gives rise to both the lncRNA MIR31HG and miR-31. Previous studies have shown that miR-31 participates in several processes relevant to vascular disease, mainly limited to SMC proliferation and migration, but also endothelial repair and macrophage function.^42–45^ In contrast, lncRNA MIR31HG remains largely unexplored in vascular biology, but accumulating evidence suggests that it is a multifunctional regulator involved in, e.g., chromatin organization, cellular senescence, and inflammatory signaling, both through nuclear and cytoplasmic mechanisms.^46^ Given the central role of these processes in vascular function and integrity, lncRNA MIR31HG represents an attractive candidate transcript downstream of the 9p21.3 locus, which may play a role in the less explored EC context. Our data further support the transcript’s relevance in EC biology, exemplified by the clear flow-dependent transcriptional responsiveness of lncRNA MIR31HG but not miR-31.

Endothelial activation is a hallmark of early vascular disease and a critical determinant of its development and progression. Silencing of *MIR31HG* in ECs induced a transcriptional program characterized by a low proliferation rate, specialized immune and metabolomic changes, suggesting that *MIR31HG* contributes to maintaining endothelial quiescence. Similarly, activation of EMT-associated processes indicates that a loss of *MIR31HG* promotes a maladaptive endothelial state signified by impaired barrier function and endothelial identity, consequently contributing to vascular remodeling. Given that lncRNA MIR31HG is flow-responsive and down-regulated under disturbed flow conditions, altered hemodynamic forces may further potentiate these processes in athero-prone vascular regions.

The association between E2F-, ETS-, and CUX/ONECUT-related regulatory activity and the transcriptional response to *MIR31HG* depletion provides candidate downstream pathways, but MARA represents an inference from sequence and expression patterns rather than direct evidence of transcription factor mediation. The observed changes therefore define candidate regulatory programs for future mechanistic studies.

Finally, we wanted to understand the genetic mechanisms underlying the 9p21.3 locus. Previous studies showed controversial results of reporter assays.^47,48^ We used the deep-learning model AlphaGenome to predict the regulatory effects on the variants in the 9p21.3 locus. While predictive models are not perfect and may highlight different parts of the same system, which is commonly described as a complementary evidence stream, we were able to converge the AlphaGenome predictions and eQTL associations in the prioritization of a putative causal variant, further supported by concordant TF binding affinity results. Thus, while remaining a putative variant, several layers of evidence suggest the high confidence of the prioritization strategy.

The CVD risk allele was associated with increased *MIR31HG* expression, while knockdown of *MIR31HG* promoted processes related to endothelial activation. Although this may seem counterintuitive, recent studies have shown that the function of *MIR31HG* is highly dependent on subcellular location rather than expression levels alone.^49^ In proliferating cells, *MIR31HG* transcripts localize both to the nucleus and cytoplasm, where its nuclear fraction interacts with Polycomb repressive complexes to regulate chromatin states and repress CDKN2A/p16INK4A expression. In contrast, during cellular senescence, *MIR31HG* is strongly induced and translocates to the cytoplasm, where it interacts with YBX1 to modulate IL1A translation. Thus, increased *MIR31HG* does not necessarily mean endothelial protection if, at the same time, it promotes RNA export to the cytoplasm. Consequently, the CVD-associated increase in *MIR31HG* may be a compensatory or context-dependent response rather than a straightforward gain of protective function. Future studies investigating the *MIR31HG* subcellular localization across different genotypes are, however, warranted to determine whether the risk locus influences *MIR31HG* function through changes in RNA export or expression levels alone.

To further validate the regulatory region, we successfully performed CRISPR activation of a candidate enhancer that markedly increased *MIR31HG* expression. This provided direct functional evidence that regulatory elements at 9p21.3 can influence *MIR31HG* transcription even through long-range chromatin interactions.

### Limitations

Despite proposing a much more complex regulatory network for the same complex 9p21.3 locus, our cohorts have some limitations. First, only individuals with ascending aortic dilatation were included, as cells could not be isolated from the small biopsies of non-dilated aortas. Second, as promoter capture Hi-C requires approximately one million cells per sample, even dilated biopsies yielded insufficient material, necessitating *in vitro* expansion of cells under static conditions. Together with the limited sample size and diverse clinical associations of the 9p21.3 locus, these conditions limit our ability to capture all the possible enhancer-promoter communication events.

## Conclusions

Taken together, our study identifies lncRNA MIR31HG as a previously unrecognized target of the 9p21.3 susceptibility locus. By integrating chromatin interaction mapping, enhancer perturbation, and functional transcriptomic analyses, we demonstrate that *MIR31HG* is regulated via long-range enhancer-promoter interactions and acts as a regulator of endothelial homeostasis by shifting the transcriptional program of ECs toward endothelial activation. These findings provide a mechanistic framework linking genetic variation at the 9p21.3 locus to endothelial dysfunction and propose *MIR31HG* as a potential mediator.

## Data Availability

The sensitive sequencing datasets are not publicly deposited but are available from the corresponding author upon reasonable request. Intermediate anonymized data/results required to replicate the analysis will be shared through Supplementary Tables upon publication.

## Contributions

Conceptualization: P.Sah., H.M.B., P.E., A.Z. Methodology: P.Sah., A.Z., H.M.B., P.E., I.V.K. Software: A.Z., N.G., A.B., O.B., S.P. Validation: A.Z., H.L., M.L. Formal analysis: A.Z., N.G., A.B., O.B., S.P. Investigation: A.Z., F-A.P., H.L., H.M.B., M.L. Resources: A.F-C., H.M.B., P.E., P.Sah., I.V.K. Data curation: A.Z. and P.Sah. Writing – original draft: A.Z., F-A.P. Writing – review and editing: A.Z., H.M.B., P.S., I.V.K., P.E., F-A.P., A.B., N.G., H.L., P.Sw., A.F-C., O.B., M.L., S.P. Visualization: A.Z., N.G., A.B. Supervision: H.M.B., P.Sah., I.V.K., P.E and P.Sw. Project administration: A.Z., H.M.B., and P.Sah. Funding acquisition: H.M.B. and P.Sah.

## Acknowledgement

The authors acknowledge support from the National Genomics Infrastructure in Stockholm, funded by Science for Life Laboratory, the Knut and Alice Wallenberg Foundation, and the Swedish Research Council, and SNIC/Uppsala Multidisciplinary Center for Advanced Computational Science for assistance with massively parallel sequencing and access to the UPPMAX computational infrastructure. H.B. was supported by the Swedish Heart Lung Foundation grants #20240686, #20240685, and #20240180, and the donation from Mr. Fredrik Lundberg and the Schörling Foundation. P.Sah. was supported by the European Union’s Horizon 2020 Research and Innovation Program under the Marie Sklodowska-Curie Grant Agreement No 860002. I.V.K. was supported by assignment 125091010189-3. The information contained in this study reflects only the authors’ views. The European Commission and the European Research Executive Agency do not accept responsibility for any use made of the information contained therein.

---

**Figure S1.**
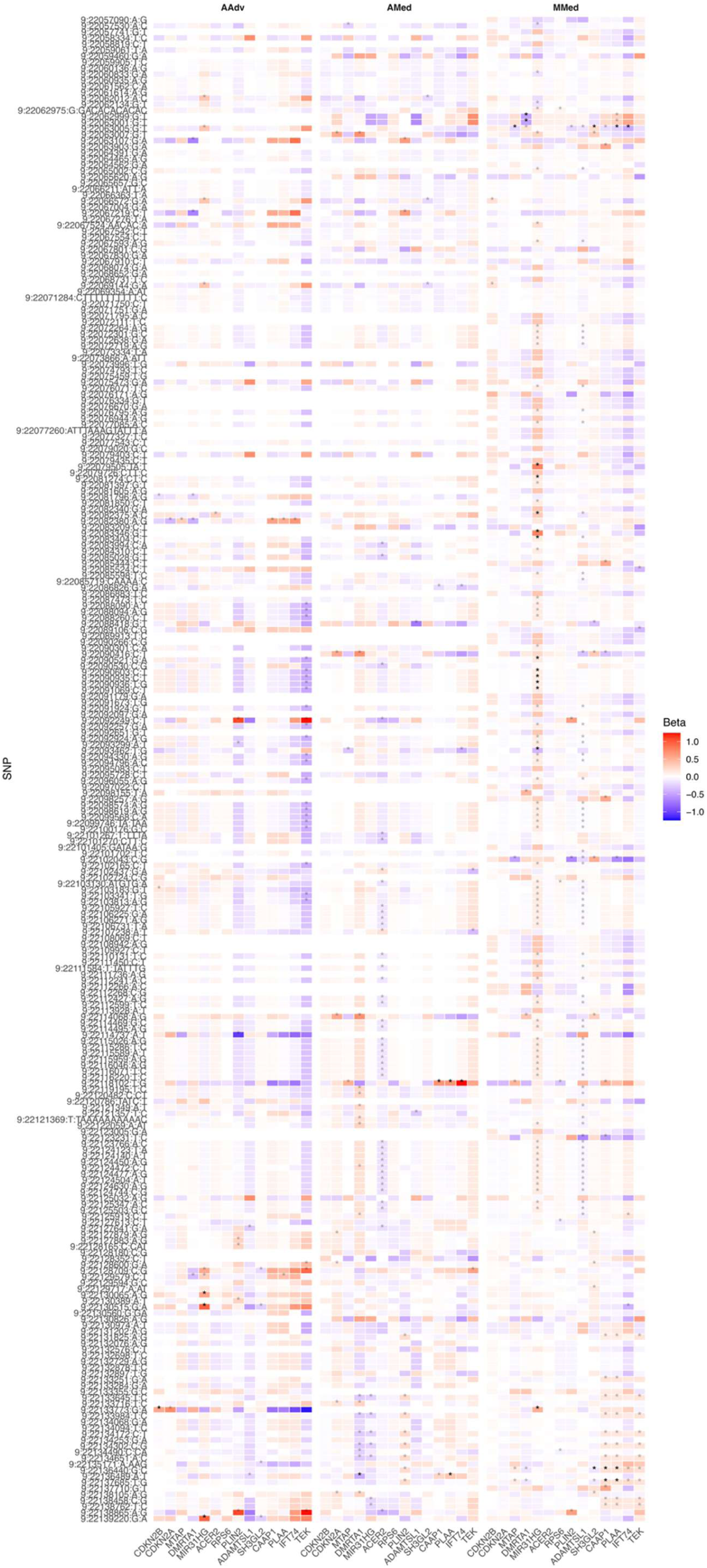
eQTL analysis of associations between genetic variation at 9p21.3 and genes interacting with the locus, as identified by HiCap. The analysis was performed in vascular tissue samples from 49 AMed, 43 AAdv, and 44 MMed biopsies. Solid stars indicate associations with an FDR < 0.05, whereas transparent stars indicate associations with *P* < 0.05 but FDR > 0.05. Color indicates the direction and magnitude of the effect size. AAdv, aortic adventitia; AMed, aortic intima-media; eQTL, expression quantitative trait locus; FDR, false discovery rate; HiCap, promoter capture Hi-C; MMed, internal thoracic (mammary) artery.

**Figure S2.**
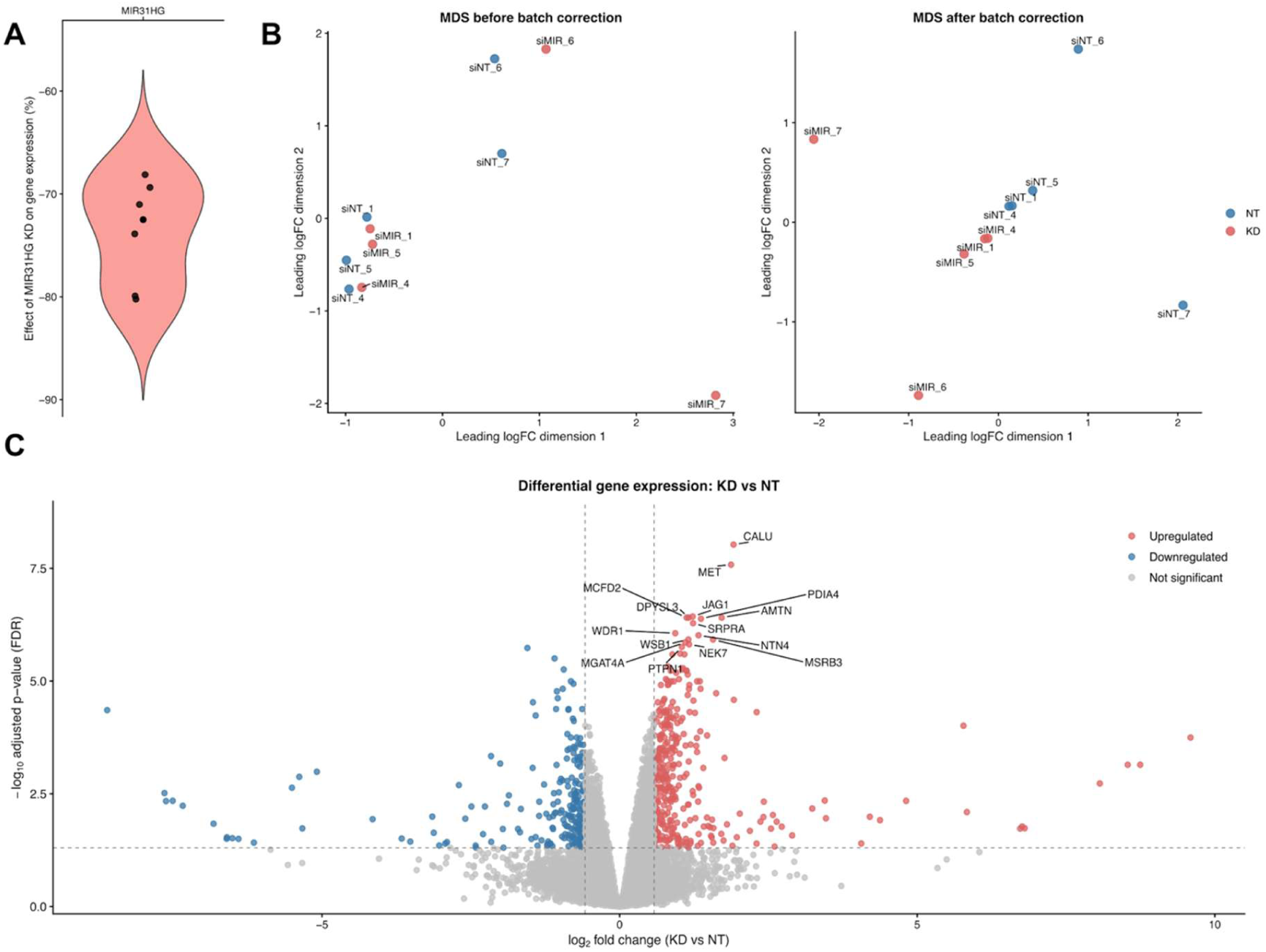
RNA-seq analysis upon *MIR31HG* KD. **A,** qPCR check of KD efficiency. **B,** MDS plots before and after the batch correction. **C,** Vulcano plot of differentially active genes. Top 15 genes are labelled. KD, knockdown; MDS, Multidimensional scaling.

**Figure S3.**
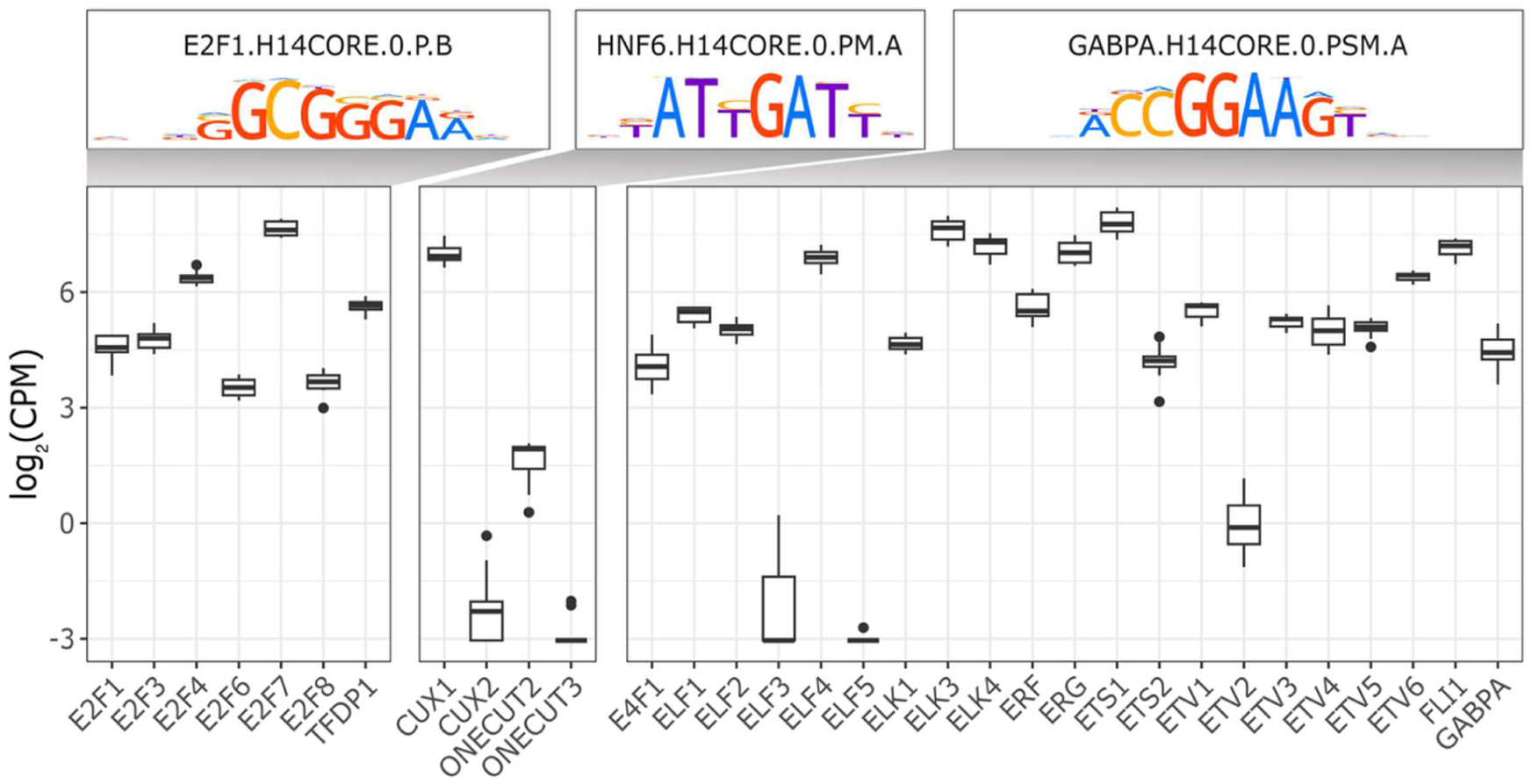
Prioritization of TFs active in endothelial cells. Members of the three prioritized transcription factor families were assessed for their expression levels in the control knockdown (KD) group. Error bars represent the standard error of the mean. CPM, counts per million; KD, knockdown; TFs, transcription factors.

## References

1. Dagenais, G. R. et al. Variations in common diseases, hospital admissions, and deaths in middle-aged adults in 21 countries from five continents (PURE): a prospective cohort study. The Lancet 395, 785–794 (2020).

2. Burton, P. R. et al. Genome-wide association study of 14,000 cases of seven common diseases and 3,000 shared controls. Nature 447, 661–678 (2007).

3. Pasmant, E. et al. Characterization of a Germ-Line Deletion, Including the Entire *INK4*/ARF Locus, in a Melanoma-Neural System Tumor Family: Identification of ANRIL, an Antisense Noncoding RNA Whose Expression Coclusters with ARF. Cancer Res. 67, 3963–3969 (2007).

4. Broadbent, H. M. et al. Susceptibility to coronary artery disease and diabetes is encoded by distinct, tightly linked SNPs in the ANRIL locus on chromosome 9p. Hum. Mol. Genet. 17, 806–814 (2008).

5. Congrains, A., Kamide, K., Ohishi, M. & Rakugi, H. ANRIL: Molecular Mechanisms and Implications in Human Health. Int. J. Mol. Sci. 14, 1278–1292 (2013).

6. McPherson, R. et al. A common allele on chromosome 9 associated with coronary heart disease. Science 316, 1488–91 (2007).

7. Helgadottir, A. et al. A Common Variant on Chromosome 9p21 Affects the Risk of Myocardial Infarction. Science (1979). 316, 1491–1493 (2007).

8. Samani, N. J. et al. Genomewide Association Analysis of Coronary Artery Disease. New England Journal of Medicine 357, 443–453 (2007).

9. Visel, A. et al. Targeted deletion of the 9p21 non-coding coronary artery disease risk interval in mice. Nature 464, 409–412 (2010).

10. Folkersen, L. et al. Relationship between CAD Risk Genotype in the Chromosome 9p21 Locus and Gene Expression. Identification of Eight New ANRIL Splice Variants. PLoS One 4, e7677 (2009).

11. Audam, T. N. et al. The 9p21.3 Coronary Artery Disease Risk Locus Modulates Vascular Cell-State Transitions via Enhancer-Driven Regulation of MTAP. Preprint at 10.1101/2025.11.18.689066 (2025).

12. Zaugg, J. B. et al. Current challenges in understanding the role of enhancers in disease. Nat. Struct. Mol. Biol. 29, 1148–1158 (2022).

13. Sanyal, A., Lajoie, B. R., Jain, G. & Dekker, J. The long-range interaction landscape of gene promoters. Nature 489, 109–113 (2012).

14. Harismendy, O. et al. 9p21 DNA variants associated with coronary artery disease impair interferon-γ signalling response. Nature 470, 264–268 (2011).

15. Corradin, O. & Scacheri, P. C. Enhancer variants: evaluating functions in common disease. Genome Med. 6, 85 (2014).

16. Sahlén, P. et al. Genome-wide mapping of promoter-anchored interactions with close to single-enhancer resolution. Genome Biol. 16, (2015).

17. Zhigulev, A. et al. Rare regulatory mutations disrupt mesenchymal molecular programs driving endocardial cushion formation in bicuspid aortic valve. Nat. Commun. 17, 3587 (2026).

18. Zhigulev, A. et al. Enhancer mutations modulate the severity of chemotherapy-induced myelosuppression. Life Sci. Alliance 7, e202302244 (2024).

19. Fegraeus, K. et al. An endothelial regulatory module links blood pressure regulation with elite athletic performance. PLoS Genet. 20, e1011285 (2024).

20. Jackson, V. et al. Bicuspid aortic valve leaflet morphology in relation to aortic root morphology: a study of 300 patients undergoing open-heart surgery. European Journal of Cardio-Thoracic Surgery 10.1016/j.ejcts.2011.04.014 (2011) doi:10.1016/j.ejcts.2011.04.014.

21. Zhigulev, A. & Sahlén, P. Targeted Chromosome Conformation Capture (HiCap). in Methods in molecular biology (Clifton, N.J.) vol. 2532 75–94 (2022).

22. Vasimuddin, Md., Misra, S., Li, H. & Aluru, S. Efficient Architecture-Aware Acceleration of BWA-MEM for Multicore Systems. in 2019 IEEE International Parallel and Distributed Processing Symposium (IPDPS) 314–324 (IEEE, 2019). doi:10.1109/IPDPS.2019.00041.

23. Abdennur, N. et al. Pairtools: From sequencing data to chromosome contacts. PLoS Comput. Biol. 20, e1012164 (2024).

24. Anil, A., Spalinskas, R., Åkerborg, Ö. & Sahlén, P. HiCapTools: a software suite for probe design and proximity detection for targeted chromosome conformation capture applications. Bioinformatics 34, 675–677 (2018).

25. Folkersen, L. et al. Unraveling divergent gene expression profiles in bicuspid and tricuspid aortic valve patients with thoracic aortic dilatation: the ASAP study. Mol. Med. 17, 1365–73 (2011).

26. Björck, H. M. et al. Altered DNA methylation indicates an oscillatory flow mediated epithelial-to-mesenchymal transition signature in ascending aorta of patients with bicuspid aortic valve. Sci. Rep. 8, 2777 (2018).

27. Chen, Y., Chen, L., Lun, A. T. L., Baldoni, P. L. & Smyth, G. K. edgeR v4: powerful differential analysis of sequencing data with expanded functionality and improved support for small counts and larger datasets. Nucleic Acids Res. 53, (2025).

28. Yoshihara, M. et al. Transcriptional enhancers in human neuronal differentiation provide clues to neuronal disorders. EMBO Rep. 26, 4 (2025).

29. Jolma, A. et al. An expanded codebook of human transcription factor DNA-binding specificity. Nature 657, 275–283 (2026).

30. Meshcheryakov, G. & Buyan, A. MARADONER: Motif Activity Response Analysis Done Right. ArXiv (2026).

31. Deviatiiarov, R. M. et al. An atlas of transcribed human cardiac promoters and enhancers reveals an important role of regulatory elements in heart failure. Nature Cardiovascular Research 2, 58–75 (2023).

32. Avsec, Ž., et al. Advancing regulatory variant effect prediction with AlphaGenome. Nature 649, 1206–1218 (2026).

33. Louwe, M. C. et al. Alternative Complement Pathway in Carotid Atherosclerosis: Low Plasma Properdin Levels Associate With Long-Term Cardiovascular Mortality. J. Am. Heart Assoc. 14, e038316 (2025).

34. Jover, E. et al. CALU polymorphism A29809G affects calumenin availability involving vascular calcification. J. Mol. Cell. Cardiol. 82, 218–227 (2015).

35. Cai, Y. et al. Salt-Inducible Kinase 3 Promotes Vascular Smooth Muscle Cell Proliferation and Arterial Restenosis by Regulating AKT and PKA-CREB Signaling. Arterioscler. Thromb. Vasc. Biol. 41, 2431–2451 (2021).

36. Gschwind, A. R. et al. An encyclopedia of human enhancer–gene regulatory interactions. Nature 657, 179–190 (2026).

37. E. Vorontsov, I., V. Kulakovskiy, I., Khimulya, G., D. Nikolaeva, D. & J. Makeev, V. PERFECTOS-APE - Predicting Regulatory Functional Effect of SNPs by Approximate P-value Estimation. in Proceedings of the International Conference on Bioinformatics Models, Methods and Algorithms 102–108 (SCITEPRESS - Science and and Technology Publications, 2015). doi:10.5220/0005189301020108.

38. Kojetin, D. J. & Burris, T. P. REV-ERB and ROR nuclear receptors as drug targets. Nat. Rev. Drug Discov. 13, 197–216 (2014).

39. Karlsson, M. et al. A single–cell type transcriptomics map of human tissues. Sci. Adv. 7, (2021).

40. Kasowski, M. et al. Variation in Transcription Factor Binding Among Humans. Science (1979). 328, 232–235 (2010).

41. Kasowski, M. et al. Extensive Variation in Chromatin States Across Humans. Science (1979). 342, 750–752 (2013).

42. Huang, S. et al. MicroRNA-31 promotes arterial smooth muscle cell proliferation and migration by targeting mitofusin-2 in arteriosclerosis obliterans of the lower extremitie. Exp. Ther. Med. 10.3892/etm.2017.5453 (2017) doi:10.3892/etm.2017.5453.

43. Guerlach, S. et al. MicroRNA-31-5p is upregulated during neointima formation and influences human coronary artery smooth muscle cell functions by regulating PIAS3 and DKK1. Cardiovasc. Res. 120, (2024).

44. miR-31 Overexpression Exacerbates Atherosclerosis by Targeting NOX4 in apoE-/-Mice. Clin. Lab. 61, (2015).

45. Yang, K. et al. Prevention of aortic dissection and aneurysm via an ALDH2-mediated switch in vascular smooth muscle cell phenotype. Eur. Heart J. 41, 2442–2453 (2020).

46. Montes, M. et al. The long non-coding RNA MIR31HG regulates the senescence associated secretory phenotype. Nat. Commun. 12, 2459 (2021).

47. Zhu, B. et al. A functional variant rs1537373 in 9p21.3 region is associated with pancreatic cancer risk. Mol. Carcinog. 58, 760–766 (2019).

48. Ali, M. W. et al. A Functional Variant on 9p21.3 Related to Glioma Risk Affects Enhancer Activity and Modulates Expression of CDKN2B-AS1. Hum. Mutat. 42, 1208–1214 (2021).

49. Montes, M. et al. The lncRNA MIR31HG regulates p16INK4A expression to modulate senescence. Nat. Commun. 6, 6967 (2015).

